# An Augmented SEIR Model with Protective and Hospital Quarantine Dynamics for the Control of COVID-19 Spread

**DOI:** 10.1101/2021.01.08.21249467

**Authors:** G. Rohith

## Abstract

In this work, an attempt is made to analyse the dynamics of COVID-19 outbreak mathematically using a modified SEIR model with additional compartments and a nonlinear incidence rate with the help of bifurcation theory. Existence of a forward bifurcation point is presented by deriving conditions in terms of parameters for the existence of disease free and endemic equilibrium points. The significance of having two additional compartments, viz., protective and hospital quarantine compartments, is then illustrated via numerical simulations. From the analysis and results, it is observed that, by properly selecting transfer functions to place exposed and infected individuals in protective and hospital quarantine compartments, respectively, and with apt governmental action, it is possible to contain the COVID-19 spread effectively. Finally, the capability of the proposed model in predicting/representing the COVID-19 dynamics is presented by comparing with real-time data.

## 1 Introduction

The Coronavirus disease of 2019, otherwise more commonly known as COVID-19, is caused by novel SARS-CoV-2 virus, a single stranded virus that belongs to RNA coronaviridae family [1]. World health organization declared COVID-19 a global pandemic on March 11, 2019 and number of people infected by this disease is growing rapidly all around the world. In this context, researchers have been working to have a clear understanding of the COVID-19 transmission dynamics and devise control strategies to mitigate the spread.

Mathematical modeling based on dynamical equations has received relatively less attention compared to statistical methods even though they can provide more detailed mechanism for the epidemic dynamics. Right from 1760, the study of dynamics of epidemics started from 1760 by modelling smallpox dynamics and since then, it has become an important tool in understanding the transmission and control of infectious diseases [2]. In 1927, Kermack and McKendrick introduced Susceptible-Infectious-Removed (SIR) compartmental modelling approach to model the transmission of plague epidemic in India [3]. Acknowledging the success of this approach, the use of mathematical modelling based approaches for the study of infectious disease dynamics has been well sough-after.

Considering the latent state that exists for COVID-19 disease, model including an additional compartment called exposed state, called Susceptible-Exposed-Infectious-Removed (SEIR) model [4] is usually used to model COVID-19 dynamics. Literature suggest widespread use of SEIR model to study the early dynamics of COVID-19 outbreak [5–10]. Effectiveness of various mitigation strategies are also studied. In [5, 6], the COVID-19 dynamics was further generalized by introducing further sub-compartments, viz., quarantined and unquarantined, and the effect of the same on transmission dynamics was presented. In [11], the classical SEIR model was further extended to introduce delays to incorporate the incubation period in COVID-19 dynamics. Recently in [12], dynamics of SEIR model with homestead-isolation was analysed by adding an additional parameter in the incidence function. However, addition of extra compartments to address the isolation/quarantine stage could serve as a better alternative. In this regard, this work attempts to model the COVID-19 dynamics by including two additional quarantine compartments to forcibly curb the disease spread. Effect of an additional control parameter, added through the selection of nonlinear incidence rate and quarantine rate functions, has also been considered.

If one attempts to mimic the actual disease spread by choosing nonlinear rates and additional compartments, associated complexities would also increase. Analysis and understanding of such composite dynamics require use of proper and effective tools. Bifurcation analysis and continuation techniques are widely employed for deciphering the nonlinear dynamics associated with physical systems [13–17]. Bifurcation techniques were widely used in analysing the dynamics of epidemic models also [18–21]. In [18], the dynamics of SEIR model considering double exposure dynamics was studied with the help of bifurcation analysis. Van den Driessche and Watmough observed the existence of saddle-node, Hopf and Bogdanov-Takens bifurcations in SIRS model, and they used bifurcation methods for their analysis [19]. Korobeinikov analyzed the global dynamics of SIR and SIRS models with nonlinear incidence [20] and used bifurcation analysis to establish the endemic equilibrium stability.

This work proposes a Susceptible − Exposed − Protective − Infectious − Hospitalized − Removed (SEPIHR) model with protective and hospital quarantine compartments as additions to conventional SEIR model. The protective and hospital quarantine rate functions determine the potency of the added compartments. An external control input is introduced as the governmental control parameter to control the spread. To convincingly simulate the COVID-19 transmission, a non-linear incidence rate is selected. Effect of a constant and adaptive quarantine rate is also studied. Dynamic analysis of the nonlinear model incorporating all the aforesaid dynamics is performed mathematically and using bifurcation techniques. The effects of control parameter on the epidemic dynamics is then studied using both bifurcation analysis and via numerical simulations. The proposed model is then compared with actual COVID-19 data to show its adequacy.

The paper is organized as follows. Section 2 presents the proposed SEPIHR model in detail. Section 3 and 4 presents the dynamic analysis and bifurcation analysis of the proposed model.Section 5 presents the numerical simulation results along with real-time data comparison. Section 6 concludes the paper.

## 2 SEPIHR Model Description

The SEPIHR model is obtained by adding additional compartments to basic SEIR model as,

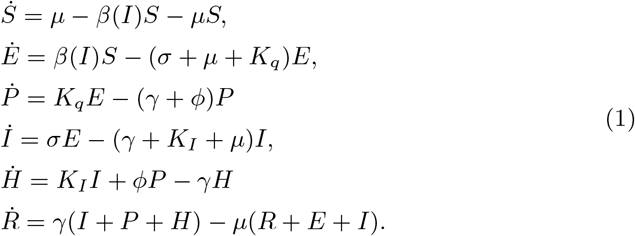

where, the state variables [*S, E, P, I, H, R*] are the fractions of total population represented in different compartments. The different compartments of the proposed model are formulated as below:

– Susceptible (*S*): The fraction of total population susceptible to the disease, but not yet infected.
– Exposed (*E*): The fraction of total population exposed to the disease, but not yet infected. They are in a latent state, after which they could show symptoms and become infective. There are chances for people in this compartment to recover without being transferred to the infective state.
– Infected (*I*): The fraction of total population who are infected and infective. After the latency period, the exposed persons are transferred to this compartment. They could be showing symptoms and mostly need hospital treatment.
– Recovered/Removed (*R*): This compartment denotes fraction of population that are either recovered from the disease or dead.
– Protective Quarantine (*P*): This is the first additional compartment in the proposed model. Since the infected compartment (*I*) dynamics is mostly governed by the fraction of exposed population, it is logical to limit the transfer from exposed to infected compartments. So, the people in the Exposed compartment (*E*) are placed under protective quarantine to limit this transmission dynamics. If the people under protective quarantine become infected, they are directly moved to hospital quarantine (second additionally added compartment, explained next), thus preventing them from being infecting others.
– Hospital Quarantine (*H*): This compartment introduces the fraction of people under treatment/quarantine in hospitals. If a person under protective quarantine is found infective (after detection tests/showing symptoms) he/she could be moved directly to hospital there by further preventing transmission.
– Birth/death rate is represented by *µ* and *γ* represents the recovery rate. Parameter *σ* is the measure of rate at which the exposed individuals become infected, in other words, 1*/σ* represents the mean latent period. Coefficients *K*_*q*_ and *K*_*I*_ represent the transfer rates of exposed individuals to protective quarantine and infected people to hospital quarantine, respectively, and *ϕ* represents the rate at which the protective quarantined people gets hospitalized. Generally, the term incidence rate or force of infection is used to model the mechanics of transmission of an epidemic. To model the complex COVID-19 transmission dynamics more precisely, a nonlinear incidence rate is used in the model.

It is normal to represent the incidence rate as a linear function of infectious class, *β*(*I*) = *β*_0_*I*. Here, *β*(*I*) represents the incidence rate and *β*_0_ denotes the per capita contact rate [18]. But, modelling the transmission dynamics/force of infection as a linear process might not be exact considering the complexities associated with it. This was first addressed in [22], where the authors used a saturated incidence rate to model cholera transmission. Since then, it became a usual practice to model the disease spread rate using nonlinear incidence functions [23]. In this paper, to represent the COVID-19 force of infection, the following nonlinear incidence rate function is used.

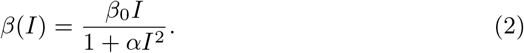

In Eq. (2), the term, *β*_0_*I* represents the bilinear force of infection and the term, 1 + *αI*^2^ represents the inhibition effect, where *α* represents the (governmental) control variable. Mathematically, Eq. (2) represents a non-monotonous function whose value increases for smaller values of *I*, and decreases for higher rates of infection (for *α* ≠ 0). This is usually interpreted as a ‘psychological’ effect, and is usually triggered via measures like isolation, quarantine, restriction of public movement, aggressive sanitation etc., [24]. It is also intuitive to model the incidence rate in this form. For instance, during initial phases, when the infection values are low, the public does not perceive the situation threatening, and the response could be frivolous, causing the disease to spread in a faster rate. As infection spreads, the public would start acknowledging the gravity of the issue and could start behaving positively to protection measures. This behavioural change is usually interpreted as a ‘psychological’ one and hence modelled as a non-monotonous function as presented in Eq.(2). In this work, *α* is represented as the percentage of total effort required to contain/mitigate the epidemic spread.

Figure 1 presents the variation of incidence rate function for different values of the (Government) control variable, *α*. For *α* = 0, indicating no governmental control, the infection could persist till the whole population is infected (Fig. 1(a)). This corresponds to bilinear incidence rate function, *β*(*I*) = *β*_0_*I*. Figures 1(b), (c) and (d) represent the incidence rate variation for different values of *α*, with magnitudes *α*_1_,*α*_2_, and *α*_3_, respectively. One could notice for *α*_1_ *< α*_2_ *< α*_3_, the incidence rate tends to fall after reaching different peak values, depending on the magnitude of *α*, signifying the importance of adequate government control in curbing the epidemic spread.

**Fig. 1.**
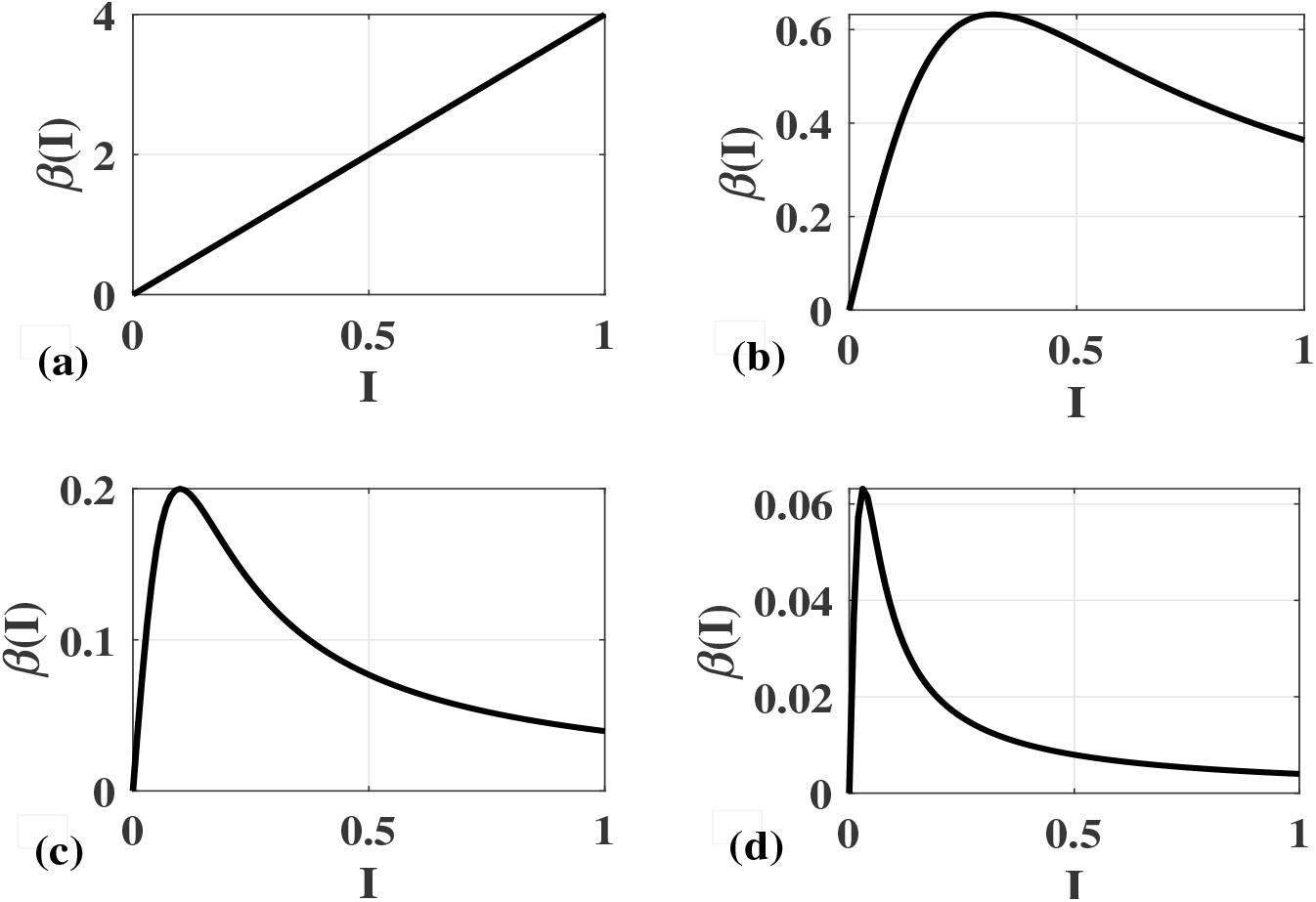
Variation of nonlinear incidence rate for different values of *α*.

A similar approach is considered for defining the protective quarantine gain *K*_*q*_. Most often, setting the quarantine rate to a constant value would not suffice when there are large number of infective individuals. This could cause an increase in exposed cases, which in turn cause a surge in infective cases. So, an adaptive mechanism to determine *K*_*q*_ value as a function of *I* could solve this problem. In this regard, *K*_*q*_ is chosen as,

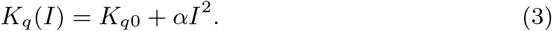

where, *K*_*q*0_ represents the static part of the function indicating the initial quarantine rate, as determined by government during the initial phase of the epidemic.

Figure 2 shows adaptive variation of *K*_*q*_ values for different magnitudes of governmental control parameters. Figure 2(a) depicts a constant *K*_*q*_ value of 0.5 for *α* = 0, or one could interpret this as a scenario where constant *K*_*q*_ value is considered, irrespective of the state of infection. Figures 1(b), (c), and (d) represent the adaptive variation of *K*_*q*_ with respect to *I* for different values of *α*. Selection of a proper *α* magnitude could depend on several other factors, and are explained in later sections.

**Fig. 2.**
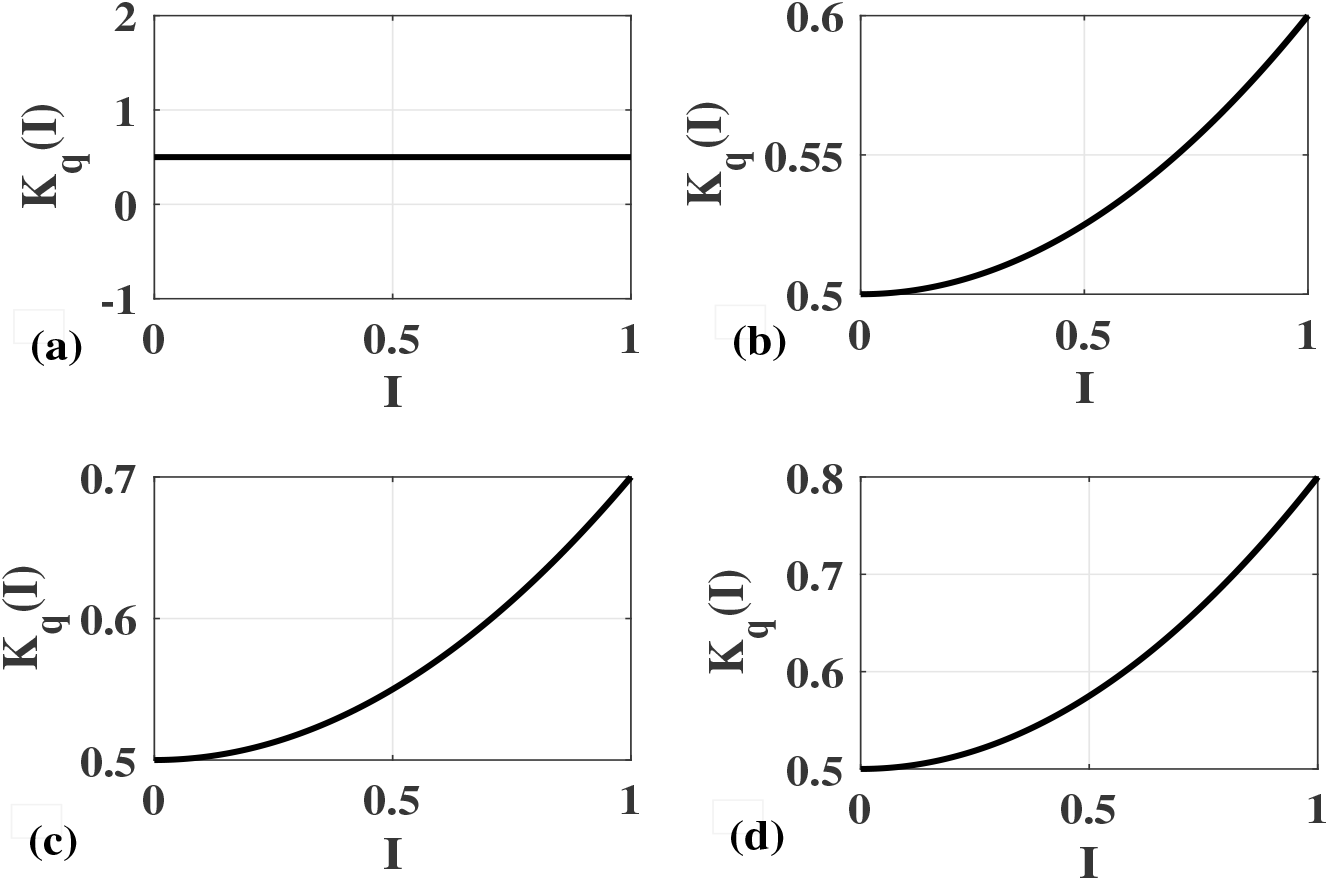
Variation of *Kq* for different values of *α*.

An overall schematic for the proposed SEPIHR model, augmenting *P* and *H* compartments along with other dependencies are explicitly demonstrated in fig. 3.

**Fig. 3.**
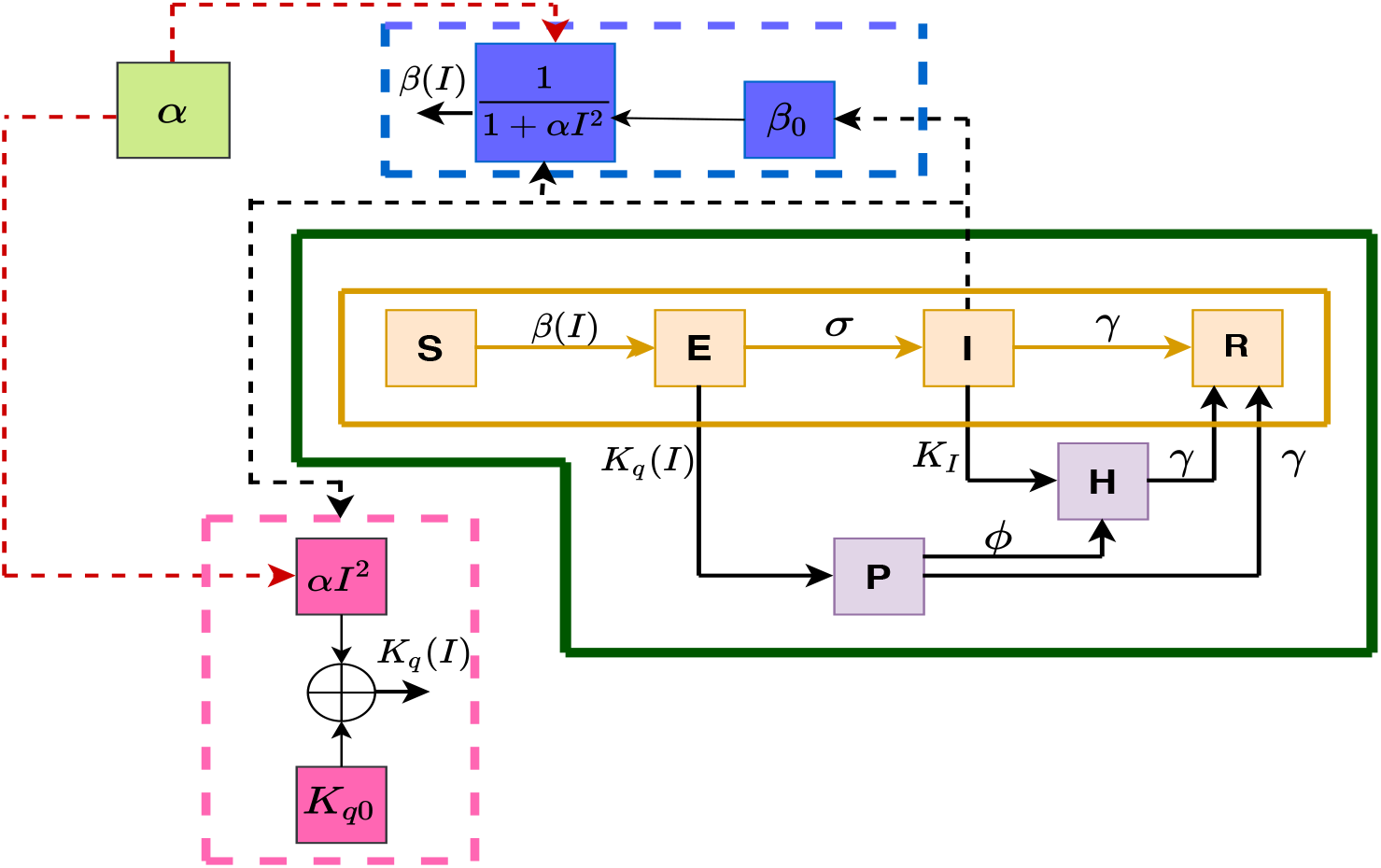
Schematic for the proposed SEPIHR model.

## 3 Dynamic Analysis

For the set of equations presented in Eq.(1), it is possible to write,

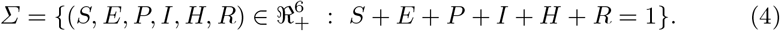

For simplicity, if one were to consider a constant value for *K*_*q*_, then the 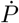 and 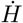 equations are now decoupled, and the system dynamics is now governed by only 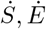 and *İ* expressions in Eq.(1). Considering this, the new set of equations can be presented as,

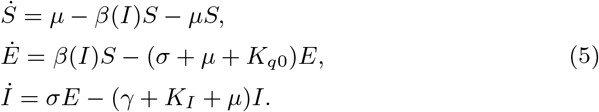

Also,

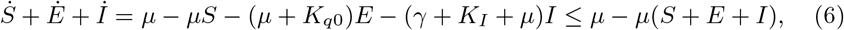

indicating the fact that, lim_*t*→∞_(*S* + *E* + *I*) ≤ 1 and the feasible region for Eq.(5) can be represented as,

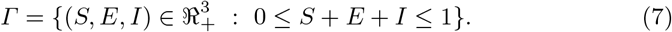

The basic reproduction number, *R*_0_, indicating the the number of secondary infections an infected individual would produce in a susceptible population, is the most important parameter that determines the epidemic outbreak. This can be obtained by solving the characteristic equation of the Jacobian of Eq.(5) about its disease free equilibrium point,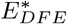 By solving Eq.(5) for 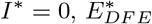 can be obtained as, (1, 0, 0). Now the Jacobian matrix for Eq.(5) is given by,

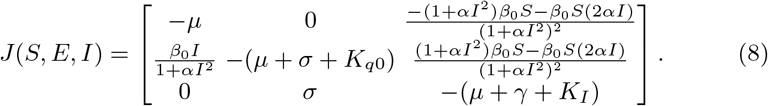

Now, to find *R*_0_, the characteristic equation |*λI* − *J* | = 0 is given as,

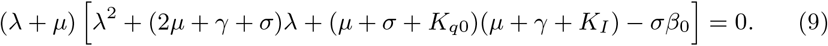

Since 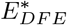 is stable, all the coefficients of Eq.(9) should be positive and all roots should have negative real parts. This implies,

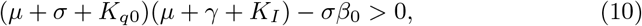

and the basic reproduction number, *R*_0_ is given by,

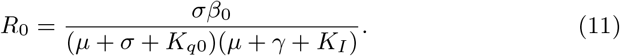

### Theorem 1

*For positive parameters, the disease free equilibrium point* 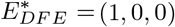 *is locally stable if R*_0_*<* 1 *and unstable if R*_0_*>* 1.

*Proof* From Eq.(8), the characteristic equation can be written as,

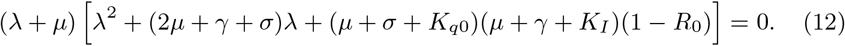

If *R*_0_*<* 1, all the coefficients of the characteristic equation are positive and all three eigenvalues are negative, indicating a stable equilibrium. For *R*_0_*>* 1, there exist a positive eigenvalue for Eq.(12) and the equilibrium solution is unstable.

### Theorem 2

*For positive parameters, there exist an endemic equilibrium* (*S*^∗^, *E*^∗^, *I*^∗^) *for R*_0_*>* 1 *and no unique endemic equilibrium for R*_0_*<* 1.

*Proof* To find the endemic equilibrium (*S*^∗^, *E*^∗^, *I*^∗^), system presented in Eq.(5) is equated to zero,

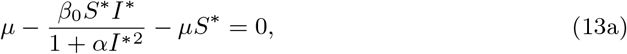

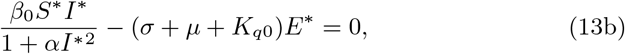

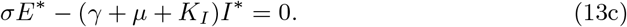

Now, from Eq.(13c),

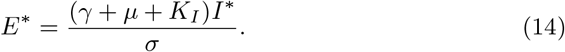

Substituting *E*^∗^ in Eq.(13b),

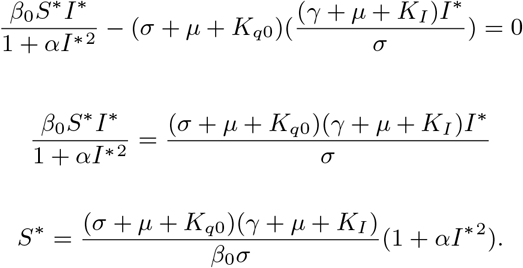

Now, *S*^∗^ can be represented in terms of basic reproduction number as,

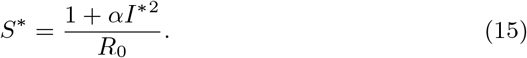

Now, one could find *I*^∗^ as the positive solution of

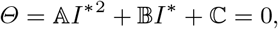

where,

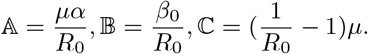

Since *µ, α*, and *R*_0_ are greater than zero, 𝔸 *>* 0 and 𝔹 *>* 0. For *R*_0_ *>* 1, ℂ *<* 0, and there exists a positive solution for *Θ*, and hence a unique endemic equilibrium. For *R*_0_ *<* 1, ℂ *>* 0 and there exists no endemic equilibrium for this condition.

From the above analysis, it is evident that the critical point for the model considered is at *R*_0_ = 1. These results are corroborated by performing the bifurcation analysis of the SEIR model presented in Eq.(1). A short introduction to the bifurcation and procedure adopted is presented next.

## 4 Bifurcation and Continuation Analysis

Through bifurcation analysis and continuation methodology, it is possible to compute all possible steady states of a parameterized nonlinear dynamical system (as function of a bifurcation parameter) along with local stability information of the steady states. Bifurcation diagrams present the qualitative global dynamics of non-linear systems. In order to perform the bifurcation analysis, the set of nonlinear ordinary differential equations of the form [25]:

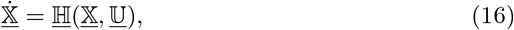

are considered, where, <u>𝕏</u> and <u>𝕌</u> are the state vector (<u>𝕏</u> ∈ ℜ^*n*^) and the control vector (<u>𝕌</u> ∈ ℜ^*m*^), respectively, and function <u>ℍ</u>(<u>𝕏</u>, <u>𝕌</u>) defines the mapping such that, ℜ^*n*^ × ℜ^*m*^ → ℜ^*n*^.

Bifurcation parameter is chosen to be varying step-wise while fixing other parameters to their constant values. At each step, fixed points are computed by solving

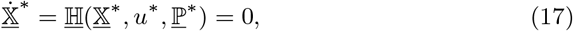

where, *u* ∈ <u>𝕌</u> and <u>ℙ</u> ∈ <u>𝕌</u> represent the bifurcation parameter and the set of fixed parameters, respectively. Once a fixed point ((<u>𝕏</u>^∗^, *u*^∗^)) is known, in a continuation, the next point (<u>𝕏</u>_1_, *u*_1_) is predicted by solving:

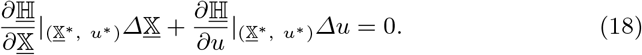

Correction step is then performed to satisfy Eq. (17) to get the next fixed point 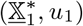. It is also possible to compute the eigenvalues (from Jacobian matrix) to determine the stability and these stability information are usually marked in the bifurcation curve.

## 5 Numerical Simulations

To corroborate the analysis presented above, bifurcation analysis is performed on the proposed model. The basic reproduction number, *R*_0_ is chosen as the bifurcation parameter to perform the analysis. The bifurcation plots presented in fig. 4 also corroborate the value of bifurcation point as *R*_0_ = 1. In order to conduct the analysis, the parameter values corresponding to COVID-19 are adapted from the literature [26–30]. For *R*_0_ *<* 1, the disease free equilibrium point is stable for all values of *R*_0_ (fig.4). For *R*_0_ *>* 1, the disease free equilibrium losses its stability and a stable equilibrium solution branch emerges, indicating endemic equilibrium solutions.

**Fig. 4.**
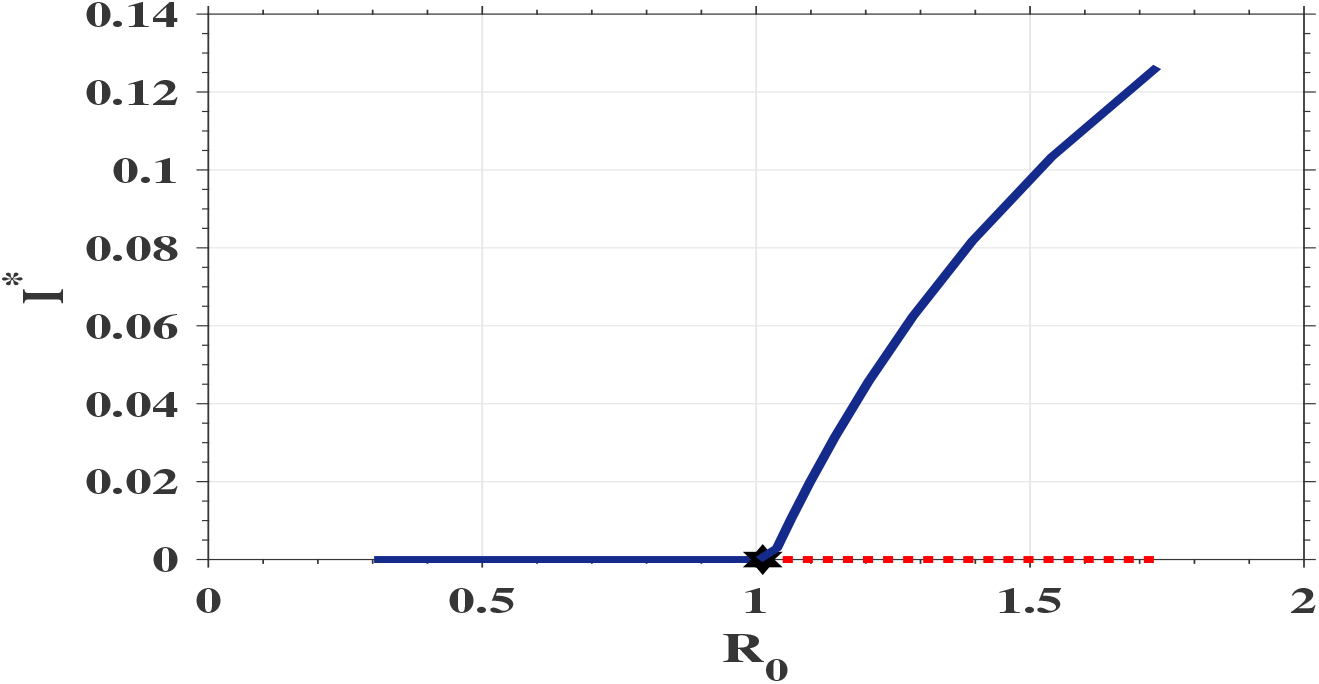
Bifurcation diagram of *I*^∗^ versus *R*_0_ — for *µ* = 0.1, *σ* = 1*/*5, *γ* = 1*/*5, *K*_*q*_ = 0, *K*_*I*_ = 0 for *α* = 0 (solid lines—stable trims; dashed lines—unstable trims; hexagram - bifurcation point).

When control parameter *α* = 0, bifurcation occurs at exactly at *R*_0_ = 1 (fig.4). But, if *α >* 0, the non-monotonicity in the incidence function dominates the dynamics and pushes the bifurcation point further towards the right, indicating more stability for the disease free equilibrium branch and a delayed outbreak, as presented in fig.5. This indicates the fact that by proper selection of *α*, it is possible to prevent the a breakout even for *R*_0_ *>* 1 scenario. This is corroborated using numerical simulation results as shown in figs. 6 and 7.

**Fig. 5.**
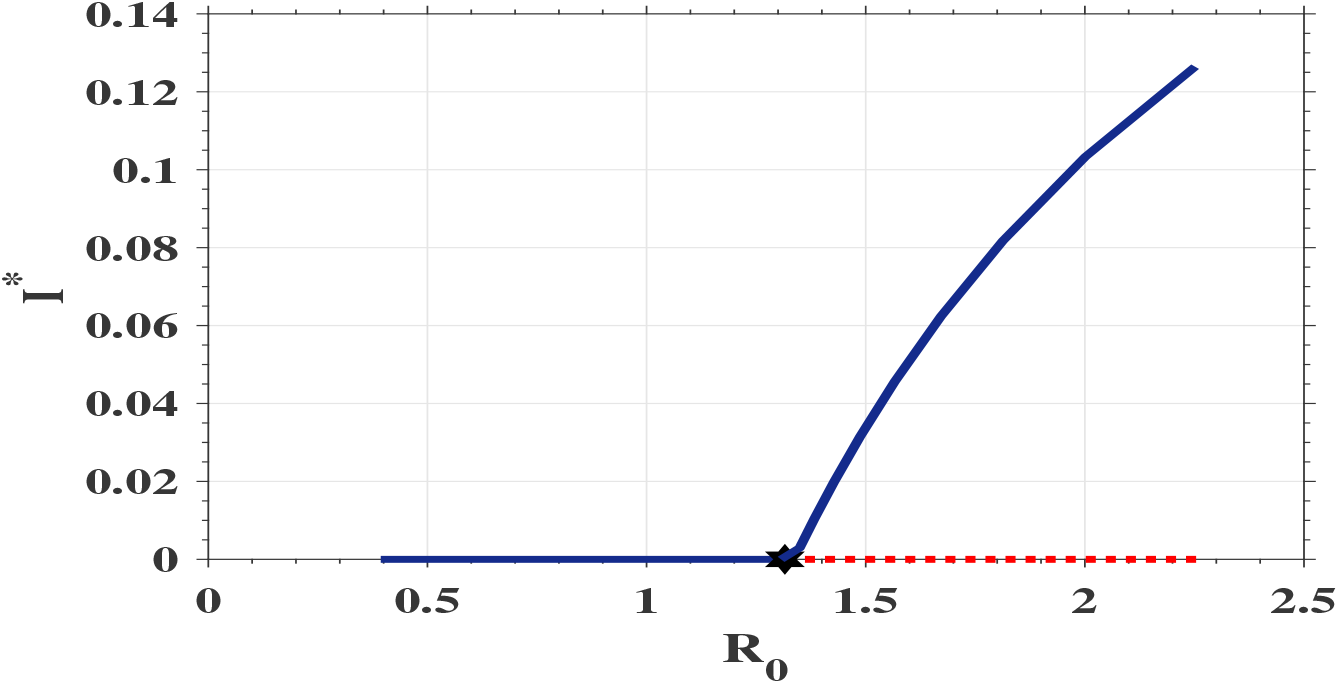
Bifurcation diagram of *I*^∗^ versus *R*_0_ — for *µ* = 0.1, *σ* = 1*/*7, *γ* = 1*/*5, *K*_*q*_ = 0, *K*_*I*_ = 0 for *α >* 0 (solid lines—stable trims; dashed lines—unstable trims; hexagram – bifurcation point).

**Fig. 6.**
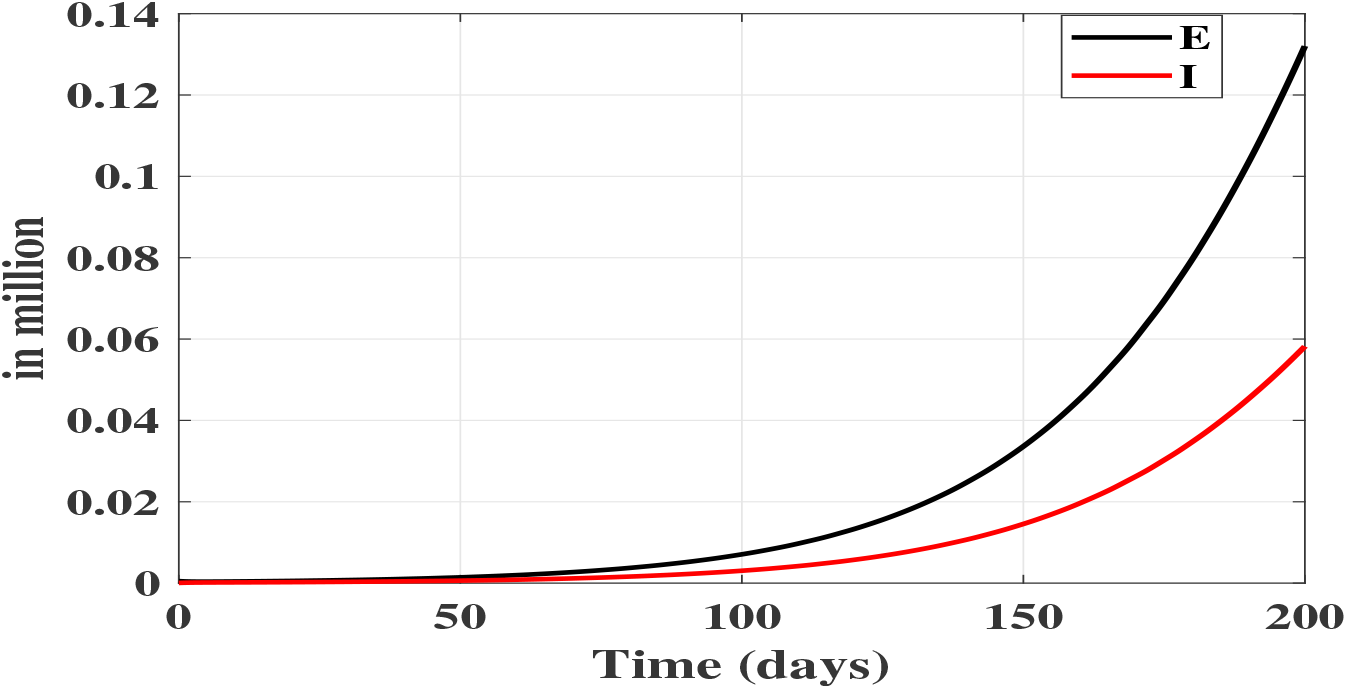
Numerical simulation results at *R*_0_ = 1.25 for fig.4 (for *α* = 0).

**Fig. 7.**
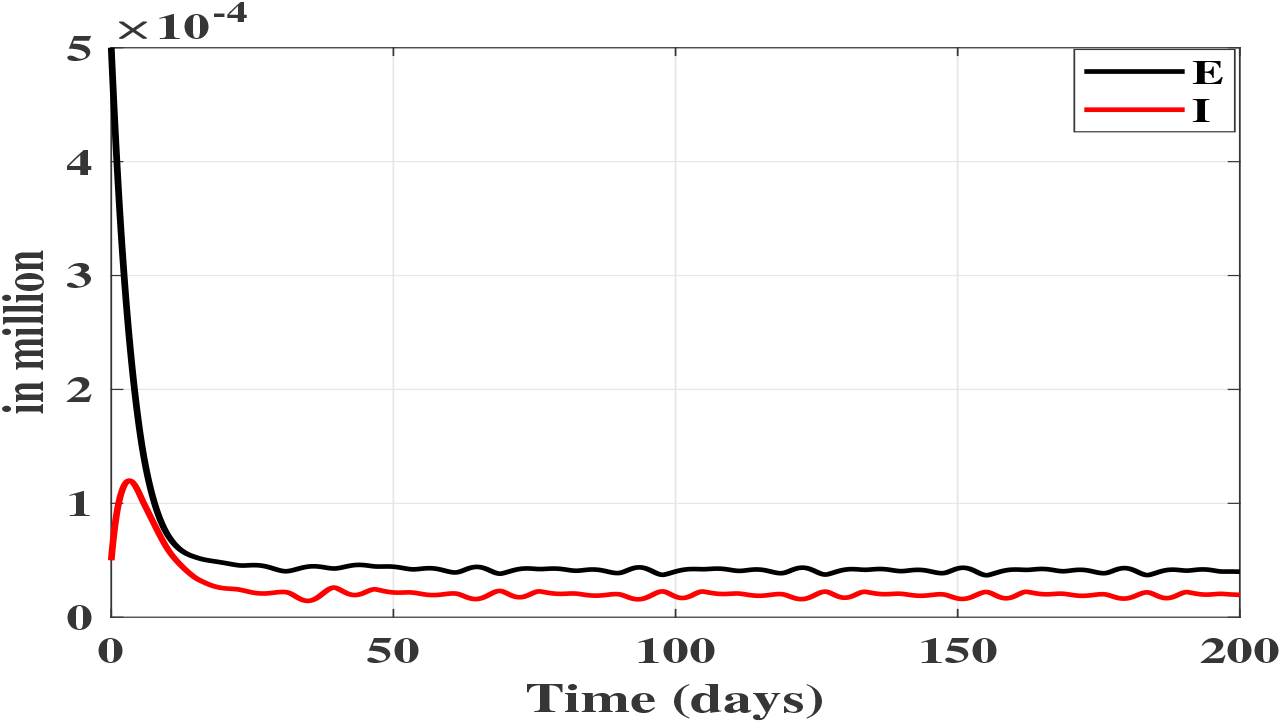
Numerical simulation results at *R*_0_ = 1.25 for fig.5 (for *α >* 0).

In order to conduct the time simulation, a city with 5 million population, out of which 90% susceptible to COVID-19 and 500 individuals exposed to the virus is considered. From fig.6, it is evident that for *R*_0_ = 1.25, endemic equilibrium exist for *α* = 0, and the stable equilibrium value corresponds to that of presented in fig.4. Since *R*_0_ value is less, it takes more time for the curves to settle to their equilibrium values, same as those suggested by the bifurcation plots. From figure fig.5, the bifurcation happens at *R*_0_ = 1.3, and for *R*_0_ = 1.25, there exist a stable disease free equilibrium solution. This could be verified from fig.7. Starting from the aforementioned initial condition, both the exposed and infected levels fall to near-zero values indicating a disease free condition. The interesting point to note here is the fact that this happens at *R*_0_ = 1.25, which usually represents an endemic state, as suggested by fig.6.

The results presented above are obtained for *K*_*q*_ = 0 and *K*_*I*_ = 0, indicating the classical SEIR model. For this set of values, *R*_0_ = 1 corresponds to a transmission rate of *β*_0_ = 0.45 (calculated using Eq.(11)). For nonzero values of *K*_*q*_ and *K*_*I*_, the two quarantine compartments become active and this affect the disease spread significantly. From Eq.(11), one could easily notice that in order to have *R*_0_ = 1, for non zero *K*_*q*_ and *K*_*I*_ values, *β*_0_ magnitude should be much higher compared to the previous case. This can be interpreted in two distinct ways. 1). For same transmission rate magnitude, the basic reproduction number will be less than that of classical SEIR model without quarantine compartments and this can reduce/stop the disease spread, depending on the value of *R*_0_. 2). With regard to SEPIHR model, it would require a higher transmission rate to sustain the disease spread compared to classical SEIR model without quarantine stages.

Figure 8 presents different *β*_0_ values required to have an *R*_0_ value of 1, to force an outbreak for different values of *K*_*q*_ and *K*_*I*_. For this set of results, the magnitude of *K*_*q*_ is fixed at different constant values rather than that of presented by Eq.(3). *K*_*I*_ values are varied in steps of 0.1 to analyze the effect. Plots are generated such that *β*_0_ values are calculated using Eq.(11) for *R*_0_ = 1 and plotted in fig. 8. One can clearly notice as values of *K*_*q*_ and *K*_*I*_ increase, the magnitude of transmission rate increases drastically. This simply means that the ‘effort’ required to curb the disease becomes lesser, or in other words, it is easier to contain the disease than using an approach without quarantine measures. For instance, for *K*_*q*_ = 0.5 and *K*_*I*_ = 0.5, disease free equilibrium exists (*R*_0_ *<* 1) up to *β*_0_ = 4.16 as opposed to *β*_0_ = 0.45 for *K*_*q*_ = 0, *K*_*I*_ = 0. This also shows the importance of adopting proper quarantine procedures.

**Fig. 8.**
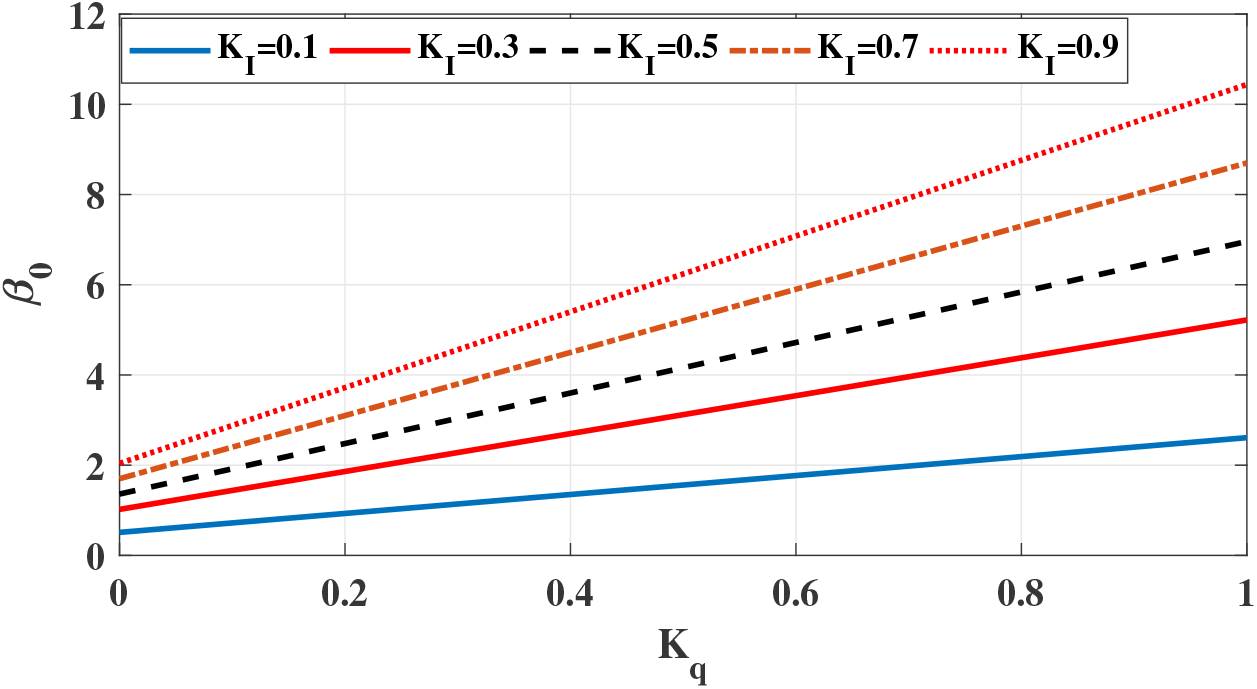
Values of *β*_0_ at different values of *K*_*q*_ and *K*_*I*_ for *R*_0_ = 1.

Figure 9 presents numerical results for *R*_0_ = 2 using the classical SEIR model. Without any quarantine measures, the number of exposed, infected and recovered cases are 1 million, 0.5 million and 0.2 million, respectively. Now, assuming the government could successfully track down and place 50% of the total exposed population in protective quarantine (*K*_*q*_ = 0.5), could hospitalize only 50% of the total number of infected persons (*K*_*I*_ = 0.5), and assuming a best case scenario of only 10% of total number of people in protective quarantine become infected (*ϕ* = 0.1), the two additional quarantine compartments in Eq.(1) become active and the number of exposed and infected cases come down to 0.32 million and 0.057 million, respectively. Number of recovered cases doubles to 0.4 million. There are 0.54 million people in protective quarantine and 0.41 million people in hospital quarantine compartments, and this additional compartments have reduced the disease spread considerably. These results are graphically presented in fig. 10, where the solid lines represent the aforementioned scenario.

**Fig. 9.**
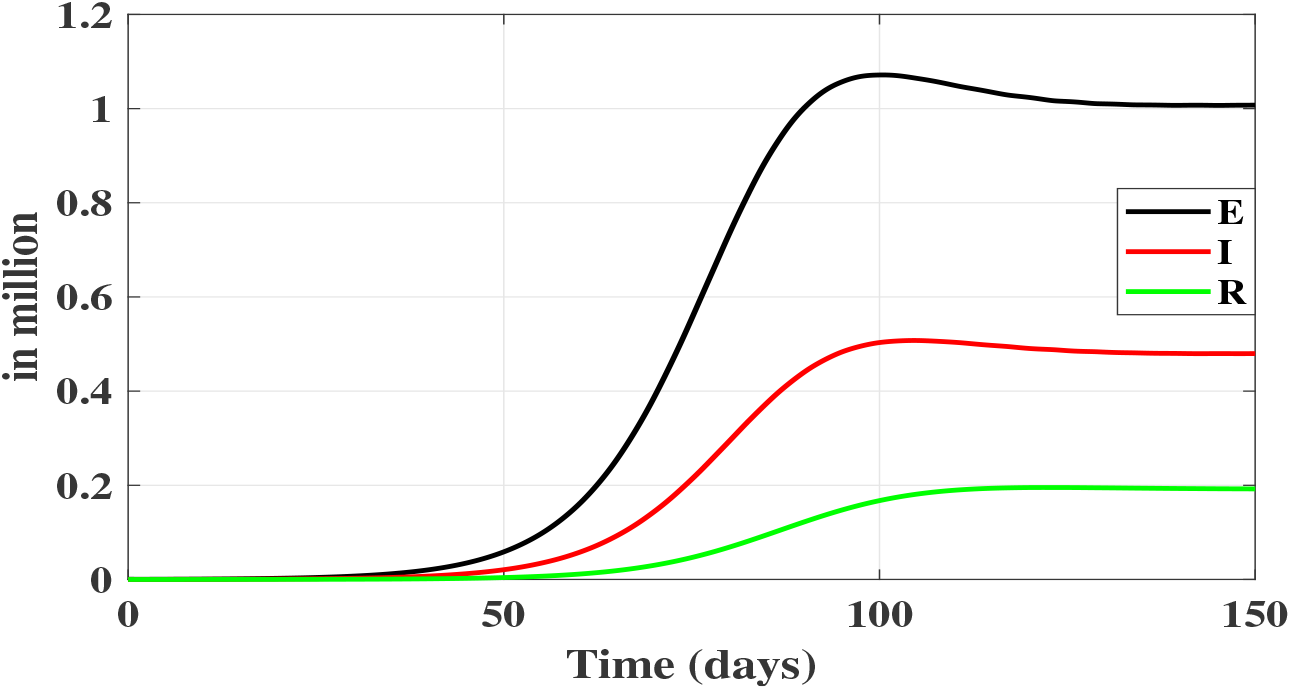
Numerical simulation results at *R*_0_ = 2 without quarantine compartments (*β*_0_ = 1, *K*_*q*_ = 0, *K*_*I*_ = 0).

**Fig. 10.**
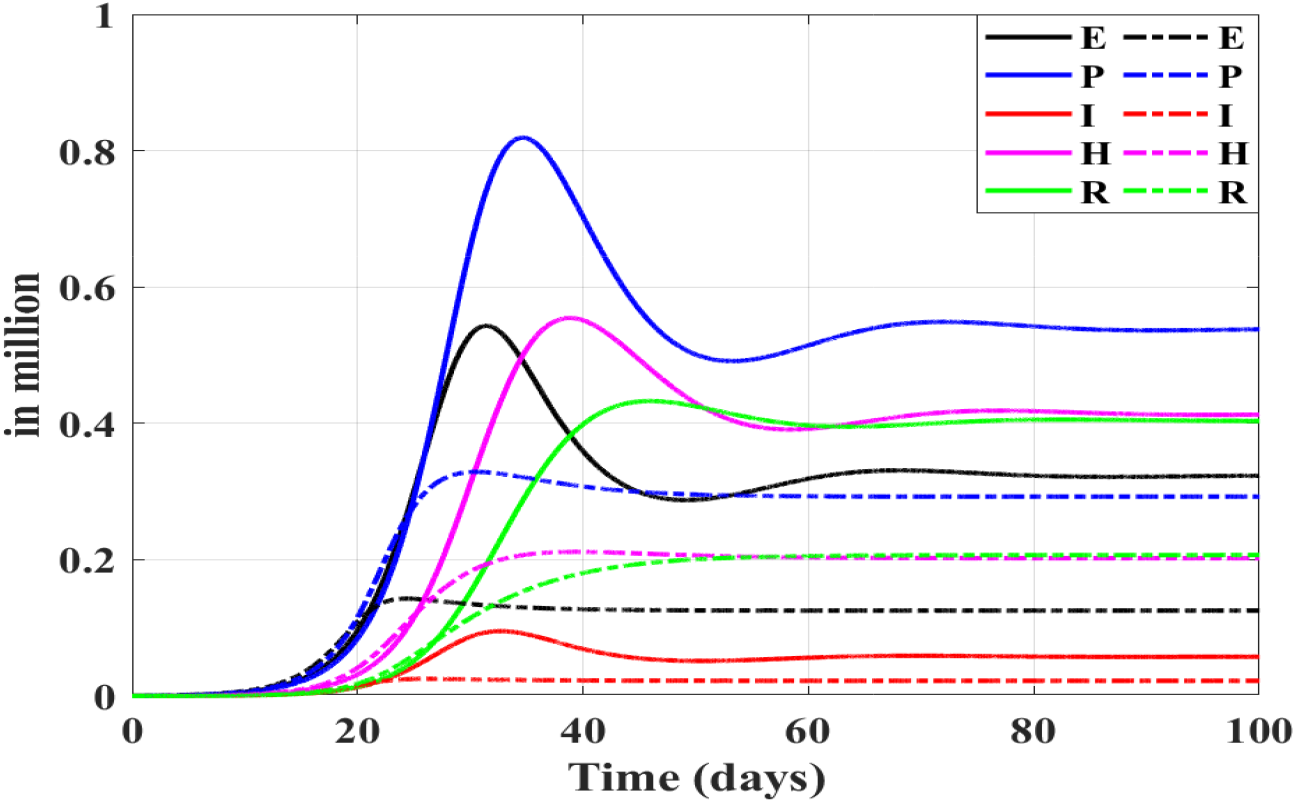
Numerical simulation results at *R*_0_ = 2 with quarantine compartments (Solid lines: for constant quarantine rate (*β*_0_ = 8.3, *K*_*q*0_ = 0.5, and *K*_*I*_ = 0.5); dotted lines adaptive quarantine rate (*β*_0_ = 8.3, *K*_*q*0_ = *K*_*q*0_ + *α* ∗ *I*^2^, *K*_*q*0_ = 0.5, and *K*_*I*_ = 0.5)).

In this work, as proposed in Eq.(3), an adaptive variation of *K*_*q*_ with respect to the rise in infections is also studied. In this regard, numerical simulations have been conducted to study the effect of such a variation in *K*_*q*_ and is presented in fig. 10 (represented as dotted lines). An initial *K*_*q*0_ value of 0.5 is chosen. As infection increases, *K*_*q*_ value also increases, depending on the value of *α* (*K*_*q*_ = *K*_*q*0_ + *αI*^2^). By using adaptive quarantine strategy, compared to constant quarantine strategy, the *E, I, P* and *H* levels to 0.125 million, 0.022 million, 0.29 million, and 0.2 million, respectively. Since *K*_*q*_ is increasing, one would expect *P* to be higher than that of previous case, and this could be true also, but for same *E* levels. But, as *K*_*q*_ increases, correspondingly *R*_0_ decreases according to the relation 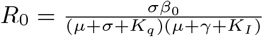. This minimizes the disease spread, lowering exposed and infected levels, causing reduced quarantine levels.

Figure 11(a) presents variation of *K*_*q*_ for *I* variation as presented in fig. 10. Curve starts from an initial value of *K*_*q*0_ = 0.5 for *I* = 0. Since *K*_*q*_ ∝ *I*, the curve follows a parabolic path, depending on the magnitude of *α*. The *K*_*q*_ value peaks around 0.75, corresponding to peak *I* value and then finally settles at 0.7. This increase in *K*_*q*_ aids in arresting the disease spread by forcing *R*_0_ value down to a smaller value. Realistically, if more people are put under protective quarantine/isolation, then the chances of disease spread come down. This is evident from the *R*_0_ plot presented in fig. 11(b). Starting from an initial value of 2, the basic reproduction number gradually drops down to a lower value of 1.55. This causes the reduction in exposed and infective levels in fig. 10 compared to a constant *K*_*q*_ scenario, where the basic reproduction number value also remains constant at *R*_0_ = 2.

**Fig. 11.**
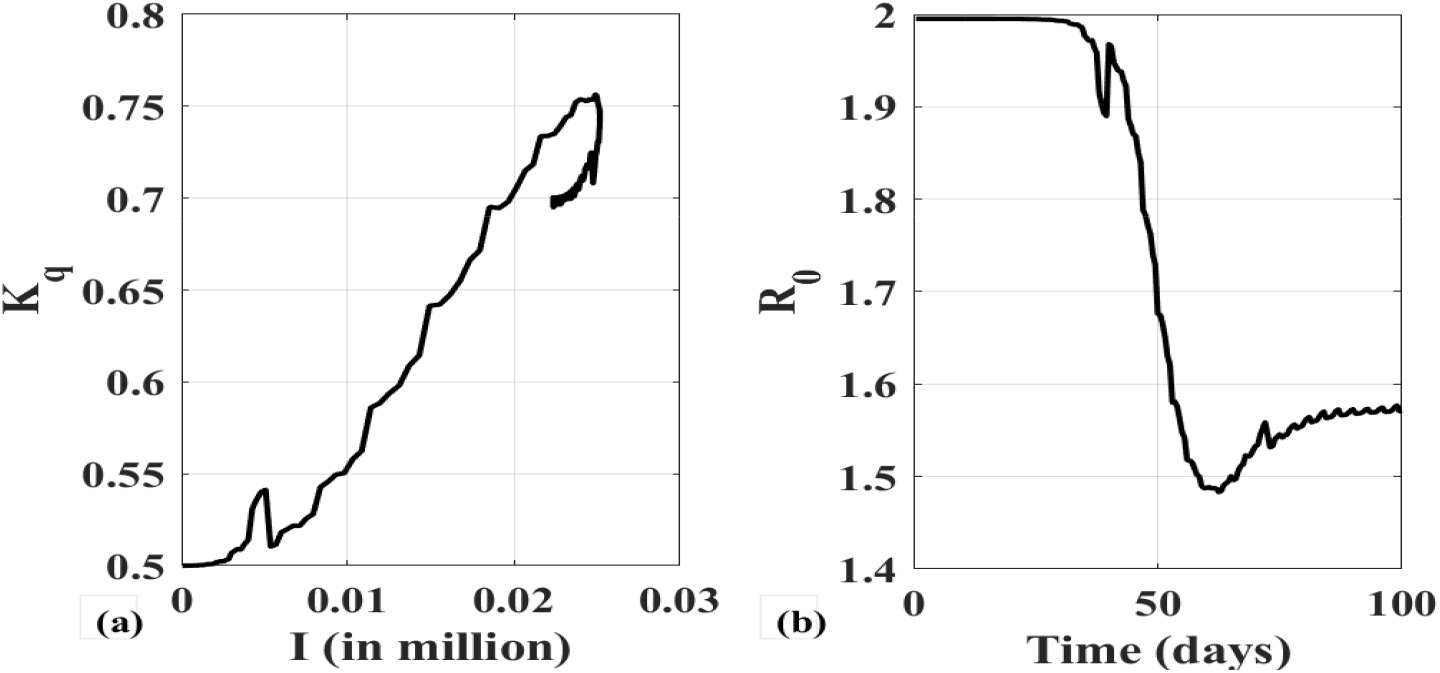
Adaptive variation of *K*_*q*_ and *R*_0_ for varying *I*, (a). variation of *K*_*q*_ with respect to *I*, (b). Change in *R*_0_ for varying *K*_*q*_.

### 5.1 Performance Evaluation of the Proposed SEPIHR Model

The efficacy of the proposed model in simulating the actual COVID-19 dynamics is verified by comparing with real-time data. Data from Kerala, one of the 28 states with 35 million population from India is considered. Kerala, famous for its ‘Kerala Model of development’ [31] is one of the developed states in India with a Human Development Index (HDI) value of 0.779 [32], highest in the country and always considered as an anomaly among developing countries. Kerala is a pioneer in implementing universal healthcare programs with a well developed healthcare system and have a literacy rate of 94% [32]. Kerala has already reached 2030 sustainable development goals in neonatal mortality rate, under five mortality rate, etc. [32]. In fact, the healthcare system is widely recognized globally, and it was named as “World’s First WHO-UNICEF Baby-Friendly State” [33].

Kerala accounts for a huge percentage of Indian diaspora and the first case of COVID-19 in India was reported in Kerala on 30/01/2020 [34]. As more and more people returned from foreign countries, the number of COVID-19 cases was on the rise. Government approached this problem via aggressive testing, contract tracing and aggressive isolation policies [35]. Efficacy of these measures helped the state in ‘flattening’ the disease curve in a much faster rate than other areas. These efforts were widely recognized globally [35–37]. They achieved this through proper contact tracing and quarantining the exposed/infected persons with the help of well developed healthcare system.

Figures 12,13,14, and 15 present the comparison of projections of *I, P, H*, and *R* states with actual data [34]. Model parameters are estimated from the data [34] and numerical simulations have been conducted to check the adequacy of the model. The estimated parameters values are given by *σ* = 1*/*7, *γ* = 1*/*12, *ϕ* = 0.125, and *µ* = 0.001. It was assumed that 80% of the exposed people are put under protective quarantine by efficient contact tracing and testing (*K*_*q*_ = 0.8) and *K*_*I*_ was estimated to be 0.45. From fig. 12, the proposed SEPIHR model predicted a peak number of 263 on 03/04/2020 compared to 262 on 05/04/2020, indicating good enough accuracy, and most importantly, the model predicted similar trend as presented by data. Figures 13 and 14 present the protective and hospital quarantine data and the trend predicted by the model. For fig. 13, like the previous case, the model predicts near accurate predictions on each days, except after 15/04/2020. After this date, the model overestimated the number of people to be under protective quarantine compared to actual data. This reduction in actual numbers could also be due to shift government policy in determining the quarantine norms.

**Fig. 12.**
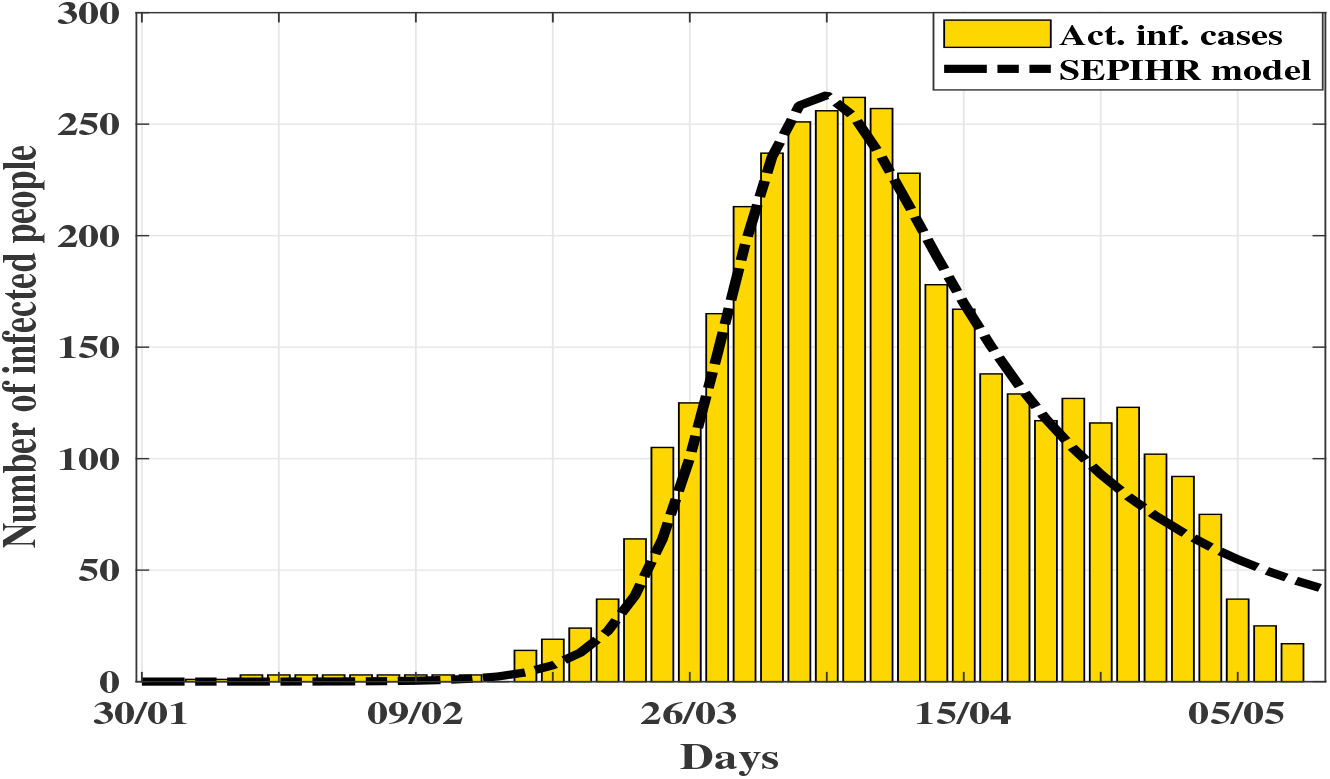
Comparison of *I* predicted using SEPIHR model with actual infected data.

**Fig. 13.**
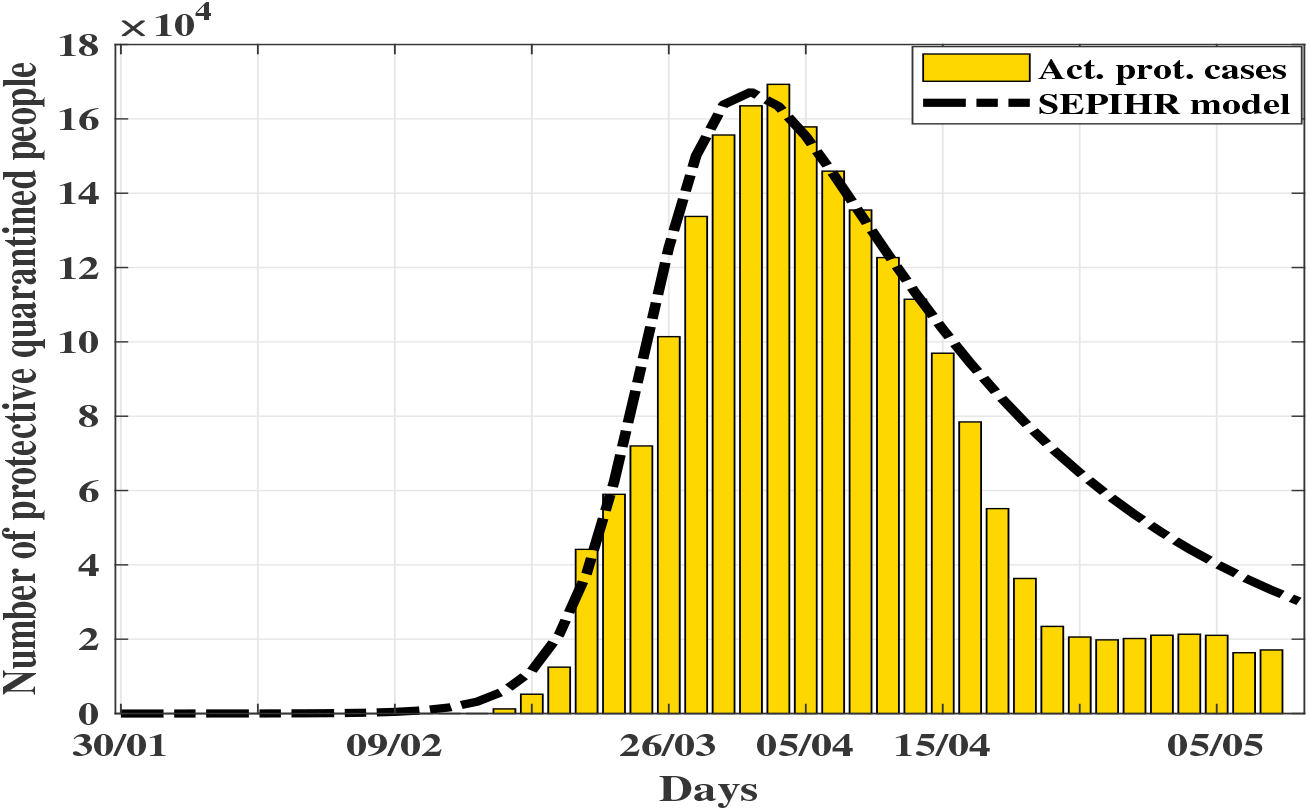
Comparison of *P* predicted using SEPIHR model with actual protective quarantined data.

**Fig. 14.**
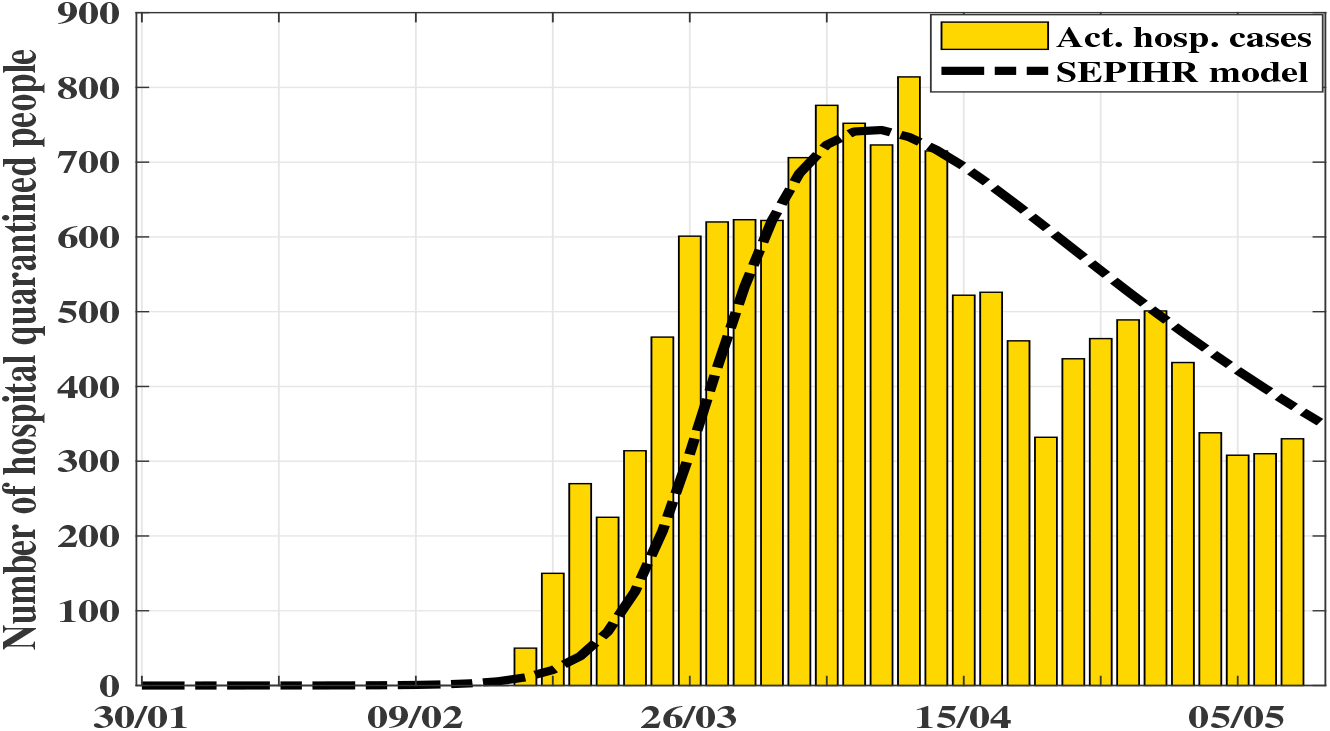
Comparison of *H* predicted using SEPIHR model with actual hospital quarantined data.

**Fig. 15.**
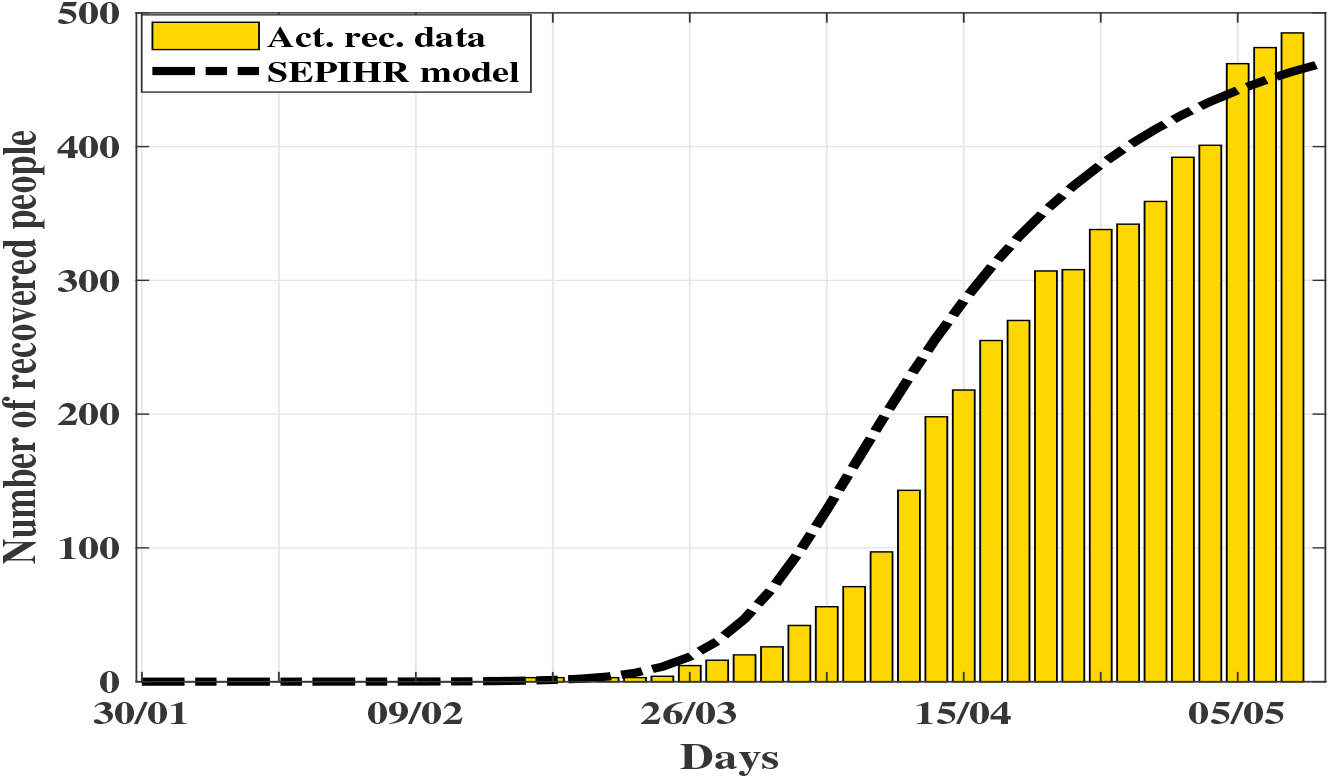
Comparison of *R* predicted using SEPIHR model with actual recovery data.

Regarding the hospital quarantine data, even though the model correctly predicts the trend, there is a mismatch in the actual predicted values (fig. 14). The model seems to be underestimating during initial phases and slightly overestimating during last phase. Again, this could be attributed to the change government norms adopted. During the initial phase of the spread, the government could have decided to place more people under hospital, fearing the spread and gradually eased the norms as things got under control. Regarding the recovery data presented in fig. 13, for the estimated parameters, even though the recovery profile overestimates the actual data by an average factor of 10%, the trend remains the same, indicating the adequacy of the proposed SEPIHR model.

## 6 Conclusions

A systematic method for the analysis and control of COVID-19 pandemic has been presented through the proposal of a new ‘SEPIHR’ model. The additional compartments, adding the dynamics of protective and hospital quarantine stages could better represent/predict the actual COVID-19 dynamics. The dynamics of government interventions in addressing the pandemic, viz., lockdown, restriction of public movement, awareness campaigns, testing, etc. is included in the model by means of nonlinear incidence function. By proper selection of *K*_*q*_, *K*_*I*_ and *α* parameters, it is possible to bring *R*_0_ below the bifurcation point or could push the bifurcation point further to higher values, thus shifting the system away from the endemic equilibrium solution branch, and preventing an outbreak. By including the protective and hospital quarantine compartments, the proposed SEPIHR model could be utilized for the prediction and performance evaluation of actual governmental quarantine efforts and could serve as a viable alternative to statistical methods in predicting and controlling the COVID-19 transmission. By comparing the predictions of the proposed SEPIHR model with actual data, the sufficiency of using a model based approach to depict/predict the COVID-19 dynamics is also emphasized.

## Data Availability

Open Source.

## Data Availability

This is necessary that we stored the data in any repository. However, our data are included in the manuscript and raw data can be released on reasonable requests.

## Declarations

### Funding

Author received no funding for this work.

### Conflict of Interest

The authors declare that they have no conflict of interest.

### Consent to participate

Not applicable.

### Consent for publication

Not applicable.

### Availability of data and material

Data is available open at [34].

### Code availability

Custom code.

## Notes

### Competing Interest Statement

The authors have declared no competing interest.

## References

1. Y. Chen, Q. Liu, D. Guo, Emerging coronaviruses: genome structure, replication, and pathogenesis, Journal of medical virology 92 (4) (2020) 418–423.

2. F. Brauer, C. Castillo-Chavez, C. Castillo-Chavez, Mathematical models in population biology and epidemiology, Vol. 2, Springer, 2012.

3. W. O. Kermack, A. G. McKendrick, A contribution to the mathematical theory of epidemics, Proceedings of the royal society of london. Series A, Containing papers of a mathematical and physical character 115 (772) (1927) 700–721.

4. H. W. Hethcote, The mathematics of infectious diseases, SIAM review 42 (4) (2000) 599– 653.

5. B. Tang, X. Wang, Q. Li, N. L. Bragazzi, S. Tang, Y. Xiao, J. Wu, Estimation of the transmission risk of the 2019-ncov and its implication for public health interventions, Journal of Clinical Medicine 9 (2) (2020) 462.

6. B. Tang, N. L. Bragazzi, Q. Li, S. Tang, Y. Xiao, J. Wu, An updated estimation of the risk of transmission of the novel coronavirus (2019-ncov), Infectious disease modelling 5 (2020) 248–255.

7. F. Binti Hamzah, C. Lau, H. Nazri, D. Ligot, G. Lee, C. Tan, et al., Coronatracker: worldwide covid-19 outbreak data analysis and prediction, Bull World Health Organ. E-pub 19.

8. S. J. Clifford, P. Klepac, K. Van Zandvoort, B. J. Quilty, R. M. Eggo, S. Flasche, C. nCoV working group, et al., Interventions targeting air travellers early in the pandemic may delay local outbreaks of sars-cov-2, medRxiv.

9. H. Xiong, H. Yan, Simulating the infected population and spread trend of 2019-ncov under different policy by eir model, Available at SSRN 3537083.

10. G. Rohith, K. Devika, Dynamics and control of covid-19 pandemic with nonlinear incidence rates, Nonlinear Dynamics (2020) 1–14.

11. Y. Chen, J. Cheng, Y. Jiang, K. Liu, A time delay dynamical model for outbreak of 2019-ncov and the parameter identification, Journal of Inverse and Ill-posed Problems 28 (2) (2020) 243–250.

12. J. Jiao, Z. Liu, S. Cai, Dynamics of an seir model with infectivity in incubation period and homestead-isolation on the susceptible, Applied Mathematics Letters (2020) 106442.

13. M. Goman, G. Zagainov, A. Khramtsovsky, Application of bifurcation methods to nonlinear flight dynamics problems, Progress in Aerospace Sciences 33 (9-10) (1997) 539–586.

14. A. Mees, L. Chua, The hopf bifurcation theorem and its applications to nonlinear oscillations in circuits and systems, IEEE Transactions on Circuits and Systems 26 (4) (1979) 235–254.

15. V. Ajjarapu, B. Lee, Bifurcation theory and its application to nonlinear dynamical phenomena in an electrical power system, IEEE Transactions on Power Systems 7 (1) (1992) 424–431.

16. S. H. Strogatz, Nonlinear dynamics and chaos: With applications to physics, biology, chemistry, and engineering, CRC press, 2018.

17. G. Rohith, N. K. Sinha, Routes to chaos in the post-stall dynamics of higher-dimensional aircraft model, Nonlinear Dynamics 100 (2) (2020) 1705–1724.

18. B. Buonomo, D. Lacitignola, On the dynamics of an seir epidemic model with a convex incidence rate, Ricerche di matematica 57 (2) (2008) 261–281.

19. P. Van den Driessche, J. Watmough, Epidemic solutions and endemic catastrophes, Dynamical systems and their applications in biology. American Mathematical Society, Providence, RI (2003) 247–257.

20. A. Korobeinikov, Lyapunov functions and global stability for sir and sirs epidemiological models with non-linear transmission, Bulletin of Mathematical biology 68 (3) (2006) 615.

21. A. Korobeinikov, Global properties of infectious disease models with nonlinear incidence, Bulletin of Mathematical Biology 69 (6) (2007) 1871–1886.

22. V. Capasso, G. Serio, A generalization of the kermack-mckendrick deterministic epidemic model, Mathematical Biosciences 42 (1-2) (1978) 43–61.

23. D. Xiao, S. Ruan, Global analysis of an epidemic model with nonmonotone incidence rate, Mathematical biosciences 208 (2) (2007) 419–429.

24. A. B. Gumel, S. Ruan, T. Day, J. Watmough, F. Brauer, P. Van den Driessche, D. Gabrielson, C. Bowman, M. E. Alexander, S. Ardal, et al., Modelling strategies for controlling sars outbreaks, Proceedings of the Royal Society of London. Series B: Biological Sciences 271 (1554) (2004) 2223–2232.

25. S. H. Strogatz, Nonlinear dynamics and chaos: with applications to physics, biology, chemistry, and engineering, CRC Press, Boca Raton, FL, 2018.

26. Q. Li, X. Guan, P. Wu, X. Wang, L. Zhou, Y. Tong, R. Ren, K. S. Leung, E. H. Lau, J. Y. Wong, et al., Early transmission dynamics in wuhan, china, of novel coronavirus–infected pneumonia, New England Journal of Medicine.

27. T. Liu, J. Hu, J. Xiao, G. He, M. Kang, Z. Rong, L. Lin, H. Zhong, Q. Huang, A. Deng, et al., Time-varying transmission dynamics of novel coronavirus pneumonia in china, bioRxiv.

28. J. A. Backer, D. Klinkenberg, J. Wallinga, Incubation period of 2019 novel coronavirus (2019-ncov) infections among travellers from wuhan, china, 20–28 january 2020, Eurosurveillance 25 (5) (2020) 2000062.

29. J. T. Wu, K. Leung, G. M. Leung, Nowcasting and forecasting the potential domestic and international spread of the 2019-ncov outbreak originating in wuhan, china: a modelling study, The Lancet 395 (10225) (2020) 689–697.

30. Q. Lin, S. Zhao, D. Gao, Y. Lou, S. Yang, S. S. Musa, M. H. Wang, Y. Cai, W. Wang,L. Yang, et al., A conceptual model for the coronavirus disease 2019 (covid-19) outbreak in wuhan, china with individual reaction and governmental action, International journal of infectious diseases 93 (2020) 211–216.

31. M. Oommen, Rethinking development: Kerala’s development experience, Vol. 2, Concept Publishing Company, 1999.

32. NITI Aayog, Healthy states, Progressive India: Report on the Ranks of States and Union Territories, NITI Aayog,(National Institution for Transforming India), Government of India, 2019.

33. Kerala named world’s first who-unicef “baby-friendly state”,- - http://www.unwire.org/unwire/20020801/28062story.asp, [Accessed : 2020 06 19].

34. Directorte of health service, government of kerala, https://dhs.kerala.gov.in/, [Accessed: 2020-06-19].

35. Aggressive testing, contact tracing, cooked meals: How the indian state of kerala flattened its coronavirus curve, https://www.washingtonpost.com/world/aggressive-testing-contact-tracing-cooked-meals-how-the-indian-state-of-keralaz-flattened-its-coronavirus-curve/2020/04/--10/3352e470-783e-11ea-a311-adb1344719a9story.html, [Accessed : 2020 06 19].

36. Coronavirus: How india’s kerala state ‘flattened the curve’, https://www.bbc.com/news/world-asia-india-52283748, [Accessed: 2020-06-19].

37. How the indian state of kerala flattened the coronavirus curve, https://www.theguardian.com/commentisfree/2020/apr/ 21/kerala-indian-state-flattened-coronavirus-curve, [Accessed: 2020-06-19].

